# BNT162b2, mRNA-1273, and Sputnik V vaccines induce comparable immune responses on a par with severe course of COVID-19

**DOI:** 10.1101/2022.02.03.21265607

**Authors:** Anna Kaznadzey, Maria Tutukina, Tatiana Bessonova, Maria Kireeva, Ilya Mazo

## Abstract

Vaccines against the severe acute respiratory syndrome coronavirus 2, which have been in urgent need and development since the beginning of 2020, are aimed to induce a prominent immune system response capable of recognizing and fighting future infection. Here we analyzed the levels of IgG antibodies against the receptor-binding domain (RBD) of the viral spike protein after the administration of three types of popular vaccines, BNT162b2, mRNA-1273, or Sputnik V, using the same ELISA assay to compare their effects. An efficient immune response was observed in the majority of cases. The obtained ranges of signal values were wide, presumably reflecting specific features of the immune system of individuals. At the same time, these ranges were comparable among the three studied vaccines. The anti-RBD IgG levels after vaccination were also similar to those in the patients with moderate/severe course of the COVID-19, and significantly higher than in the individuals with asymptomatic or light symptomatic courses of the disease. No significant correlation was observed between the levels of anti-RBD IgG and sex or age of the vaccinated individuals. The signals measured at different time points for several individuals after full Sputnik V vaccination did not have a significant tendency to lower within many weeks. The rate of neutralization of the interaction of the RBD with the ACE2 receptor after vaccination with Sputnik V was on average slightly higher than in patients with a moderate/severe course of COVID-19. The importance of the second dose administration of the two-dose Sputnik V vaccine was confirmed: while several individuals had not developed detectable levels of the anti-RBD IgG antibodies after the first dose of Sputnik V, after the second dose the antibody signal became positive for all tested individuals and raised on average 5.4 fold. Finally, we showed that people previously infected with SARS-CoV-2 developed high levels of antibodies, efficiently neutralizing interaction of RBD with ACE2 after the first dose of Sputnik V, with almost no change after the second dose.

## Introduction

In March 2020, the World Health Organization declared the disease caused by a virus from the *Coronaviridae* family (1), known as the severe acute respiratory syndrome coronavirus 2 (SARS-CoV-2), a pandemic. At the time, there were 20 thousand registered cases of SARS-CoV-2 disease (COVID-19) with less than a thousand deaths. As of October 2021, 240 million cases are reported with over 4.8 million deaths registered worldwide.

By March 19th, 2020, the global pharmaceutical industry announced a major commitment to address COVID-19. Vaccines, which have been in development since then, are intended to provide acquired immunity against SARS-CoV-2 among the world population and help conquer the pandemic. Knowledge about the structure and function of coronaviruses causing diseases like severe acute respiratory syndrome (SARS) and Middle East respiratory syndrome (MERS) (2,3) aided the accelerated development of various vaccine technologies. Currently, several COVID-19 vaccines have demonstrated efficacy as high as 95% and more during Phase III trials. As of October 2021, 6.5 billion doses of COVID-19 vaccines have been administered worldwide. The following vaccines were authorized by at least one national regulatory authority for public use: RNA vaccines (Pfizer–BioNTech BNT162b2, and Moderna mRNA-1273), conventional inactivated vaccines (Sinopharm BBIBP-CorV, Chinese Academy of Medical Sciences vaccine, CoronaVac, Covaxin, WIBP-CorV, CoviVac, COVIran Barakat, Minhai-Kangtai, and QazVac), viral vector vaccines (Oxford–AstraZeneca AZD1222, Gamaleya Sputnik V, Gamaleya Sputnik Light, Convidecia Ad5-nCoV, and Janssen-Johnson & Johnson Ad26.COV2.S), and protein subunit or peptide vaccines (EpiVacCorona, RBD-Dimer ZF2001, Abdala, and Soberana 02). Over three hundred vaccine candidates are at various stages of development.

A vaccine typically contains an agent that resembles a disease-causing microorganism. It stimulates the immune system to define the agent as a threat to recognize and destroy it in the future (4). Although COVID-19 vaccines do not offer protection in every case, it has been shown that even with breakthrough infections, the viral load in vaccinated individuals is significantly decreased (5) and the risks of transmitting the infection appear to be several times as low (6–8). One of the main immune system responses to vaccines is production of antibodies. It is thus of interest to compare the antibody development characteristics in response to vaccines of various types.

Moderna mRNA-1273 and Pfizer–BioNTech BNT162b2 are RNA vaccines composed of nucleoside-modified mRNA encoding an altered version of the spike protein of SARS-CoV-2, which is encapsulated in lipid nanoparticles (9,10). Gamaleya Sputnik V, or Gam-COVID-Vac, is a viral two-vector vaccine based on two strains of common cold human adenoviruses, Ad26 and Ad5, also containing a gene encoding the SARS-CoV-2 spike protein (11). All three vaccines require two-part administration.

The spike (S) glycoprotein in its trimeric form is in charge of the initial interaction of the virus with the receptors of the host cell. S-protein consists of two subunits, S1 and S2, separated by the furin protease cleavage site. Cleavage of the spike protein at this site facilitates viral entry into the cell, and occurs only in SARS-CoV-2, not SARS or MERS, possibly being one of the reasons for the higher infectivity of the former (12). Receptor binding domain (RBD) is a part of the S1 subunit, represented by aminoacids from 333 to 527, that binds the angiotensin converting enzyme 2 (ACE2) receptor on the cell surface. Particular amino acids directly involved in this interaction are 438-506 (RBM) (13). Several structural studies indicated that SARS-CoV-2 has stronger affinity for the ACE2 receptor than SARS (14).

The RBD is known to stimulate production of the IgG antibodies which can bind to the virus and prevent it from attaching to the human ACE2 receptors (15–18), thus anti-RBD IgG are often called neutralizing antibodies. In recent studies, it was shown that about a third of samples from individuals with previous SARS-CoV-2 infection do not seem to contain a detectable amount of the neutralizing antibodies, and that their low or absent titers positively correlate with the possibility of critical illness (patient death) (19). This allows to suggest that obtaining a high level of neutralizing antibodies is a preferable outcome of a vaccination.

In this work, we compared development of the IgG antibodies in different individuals as an immune system response after vaccination with Pfizer–BioNTech, Moderna, or Sputnik V, using a unified ELISA-based assay previously developed in our laboratory. This assay simultaneously tests for antibodies to two types of viral antigens, nucleocapsid protein (N) and RBD (20). To assess the serum neutralization capabilities in individuals vaccinated by Sputnik V, we tested the ability of the respective serum to prevent RBD from binding to the ACE2 receptor, and compared it with that in previously infected patients. We also analyzed the levels of the anti-RBD IgG antibodies developed after vaccination and previous infection with COVID-19, studying groups of patients with light/asymptomatic or moderate/severe courses of the disease. For several patients, we have performed the test multiple times after the vaccination, to reveal the tendency for the antibody levels to drop or raise over time. Finally, we assessed the levels of the anti-RBD IgG antibodies in vaccinated patients who have had COVID-19 prior to vaccination at different time points, as well as the ability of their serum to neutralize interaction with ACE2.

## Materials and Methods

### Samples

For the vaccine comparison analysis, 79 serum samples from vaccinated individuals were used; 18 samples were obtained from people vaccinated by both doses of BNT162b2 (Pfizer–BioNTech), 16 from vaccinated by both doses of mRNA-1273 (Moderna) and 58 samples from 40 people vaccinated by the Sputnik V (Gam-COVID-Vac, Gamaleya), including 18 samples taken after the first administration of the vaccine and 40 after the second (Supplementary Table S1).

To compare the levels of anti-RBD levels within the same individuals over time, additional samples from 17 Sputnik V vaccinated individuals were taken in 7-30 weeks after the vaccination.

For the comparison of vaccine-induced and disease-induced antibody development effects serum samples from 40 COVID-19 patients were used, with infection confirmed by RT-PCR, antigen tests, or alternative commercial SARS-CoV-2 antibody tests. Of these, 20 samples belonged to a group of patients with asymptomatic/light course of the disease (who had experienced only anosmia, fatigue, headache, fever below 37.5 C, or had no symptoms), and 20 samples belonged to a group with moderate/severe course of the disease (who had experienced fever above 37.5 C, cough, lung lesions, and, in several cases, were hospitalized) (Supplementary Table S1).

For the neutralization effect study 38 samples from individuals vaccinated by Sputnik V and 16 samples of previously infected individuals (8 samples from asymptomatic/light symptom group and 8 samples from moderate/severe symptom group) were used.

For the study of vaccinated individuals who previously encountered COVID-19 four additional samples were taken, with infection confirmed by RT-PCR or antigen tests.

Blood samples were taken with either venipuncture in a commercial certified laboratory, or with finger prick using 20μl Mitra Cartridge (2-Sampler, Neoteryx, USA) according to the manufacturer’s protocol.

As negative controls, 8 pre-pandemic samples were used, tested HIV- and hepatitis-negative, provided by the Laboratory of cell cultures and cell engineering of the Institute of Cell Biophysics RAS.

All procedures were approved by the Commission on Biosafety and Bioethics (Institute of Cell Biophysics – Pushchino Scientific Center for Biological Research of the Russian Academy of Sciences, Permission no. 1 of June 12, 2020) in accordance with Directive 2010/63/EU of the European Parliament. The patients/participants provided their written informed consent to participate in this study.

### Dual-antigen testing ELISA assay

The signal of anti-RBD and anti-N IgG antibodies in the samples was analyzed using the dual-antigen VirIntel assay (20). ELISA was made as described in (20) with minor modifications. In brief, the wells of the plate were filled with 98 μL of PBS-T containing 1% casein (1× Casein in PBS ready to use solution (#37528, Thermo Fisher Scientific, USA) with 0.1% TWEEN-20 added). Then, 2 μL of each sample was added to each well, and the plate was incubated for 2 h at 23 °C (RT). If a sample was collected with finger prick, then 200 μL of PBS-T containing 1% casein was added, and blood was extracted via the 1-hour incubation at 37 °C and constant shaking. Then 70 μL of the resulting sample was added to each well. After 3 washes with 300 μL of PBS-T, 100 μL of anti-human IgG HRP-conjugated secondary antibody (A01854, GenScript, USA) diluted 1:3000 in PBS-T+1% casein was added to the wells. The plate was incubated for 1 h at 23 °C (RT), washed three times with PBS-T and stained with SigmaFast OPD (P9187, Sigma, USA). The resulting absorbance was measured on a Biotek Synergy H1 plate reader (Biotek Instruments, USA) at 490 nm. Each sample was assayed in duplicate.

### ELISA result analysis

The antibody level for each individual was determined by comparison of the obtained optical density value of the respective sample to the threshold of the assay, as specified in the VirIntel test protocol (20). The ratio of the signal to the threshold for each sample is referred to as the “signal to cut-off ratio” (S/CO) and the result is considered positive if the S/CO is above 1.

Throughout the study, the S/CO for antibodies to both RBD and N antigens were measured for each sample. All three types of studied vaccines (Moderna mRNA-1273, Pfizer–BioNTech BNT162b2 and Gamaleya Sputnik V) are based on the spike protein of the SARS-CoV-2 and were expected to yield antibody production only to the RBD antigen. If the results from the assay were also positive for the N-antigen, we presumed that the respective individual had encountered COVID-19 in the past, during or after the vaccination process. These samples were excluded from further analysis unless otherwise specified within the Results and Discussion sections.

### Neutralization assay

The ability of serum to inhibit the RBD binding to ACE2 receptor was measured using the SARS-CoV-2 Surrogate Virus Neutralization Test (sVNT) Kit (ProteoGenix, France) according to the manufacturer’s protocol.

### Statistical analysis

The statistical significance calculations were done and respective P-values were obtained using the Mann-Whitney U test for comparison of S/CO value distributions between different sets of samples, and using Spearman’s correlation coefficient method for estimating the S/CO difference after first and second administrations of the vaccine in individuals within the Sputnik V vaccinated group, and for assessing correlation between individual’s age and S/CO. P-value of < 0.05 was used as a threshold for statistical significance evaluation.

## Results

### 3.1 Antibody signal for the Sputnik V vaccinated individuals after the first and the second doses

To analyze the levels of immune response after the first and the second vaccine doses, 18 Sputnik V vaccinated individuals were tested twice: shortly before second vaccination and two to three weeks after. Three tested individuals with highest levels of anti-RBD antibodies were also positive for the anti-N antibodies, which might indicate previous infection with SARS-CoV-2. One more individual had shown positive anti-N results after the second dose, presumably having been infected between the doses. After eliminating these four samples from the dataset, 14 results were left to compare and are shown on Figure 1. Of them, 11 demonstrated positive results for the anti-RBD antibodies after the first dose, with S/CO ratio ranging from 1.21 to 10.27 with an average of 3.45 and a median of 2.45. Three patients tested RBD-negative after the first dose of Sputnik. After two doses all 14 patients were tested positive.

**Fig. 1.**
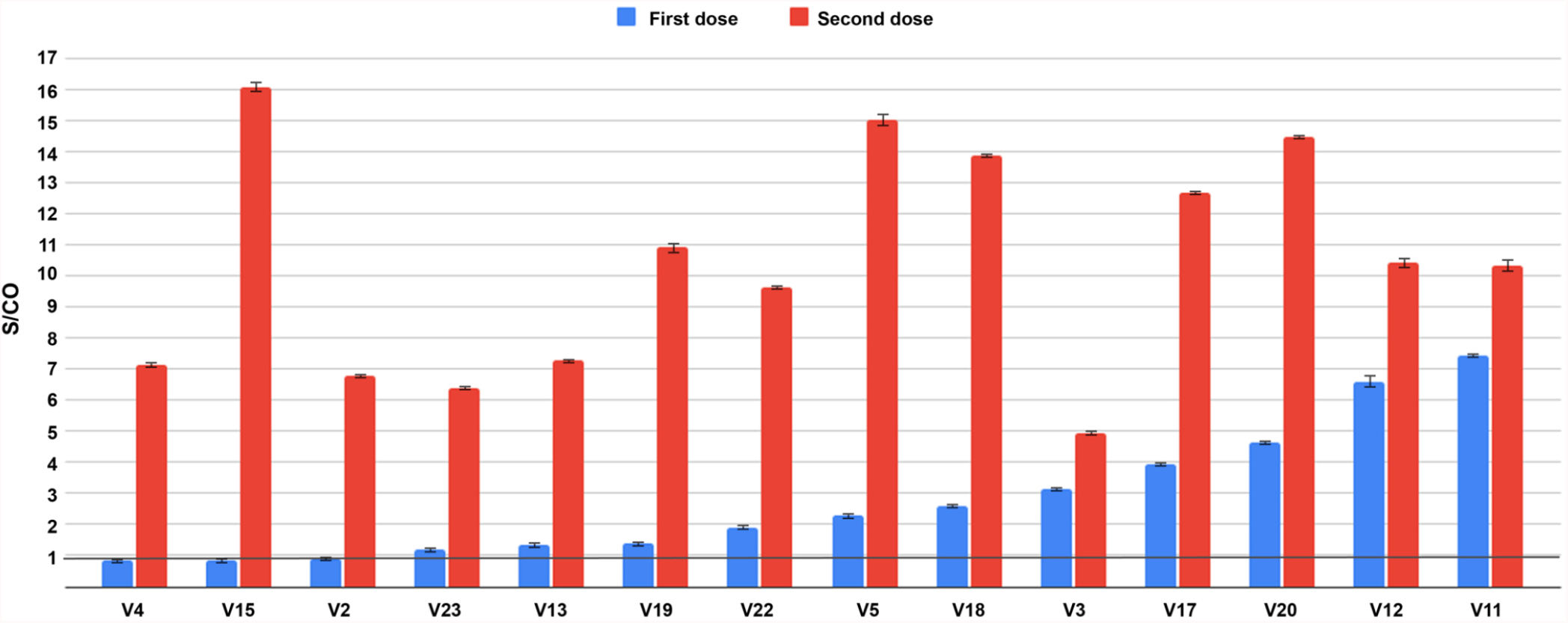
Comparison of S/CO for the anti-RBD IgG antibodies in the individuals after the first (blue) and the second (red) administration of the Sputnik V vaccine. Threshold S/CO value is 1 and is indicated with a black horizontal line.

Overall, the difference between the S/CO result distributions in samples after the first and second dose of Sputnik vaccination as measured by Mann-Whitney test was very prominent (P-value 2.67 × 10^− 05^). The S/CO after the second dose was on average 5.4 times higher (Fig. 1). At the same time, Spearman’s coefficient analysis showed that there was no significant correlation between the signal after the first and second dose within individuals (P-value 0.39); for some individuals the signal raised much more drastically (for example, for V15 on Fig. 1) than for others (V11 on Fig. 1).

### 3.2. Antibody signal for the individuals after two-dose vaccination by Sputnik V, Moderna, Pfizer-BioNtech, and previously infected individuals

In all 40 cases of the Sputnik V-vaccinated individuals tested after the second dose, positive results for the anti-RBD antibodies were demonstrated (Figs. 1 and 2). However, the S/CO ranged dramatically, with values between 1.06 and 21.0, with the average of 10.95 and median of 11.07.

**Fig. 2.**
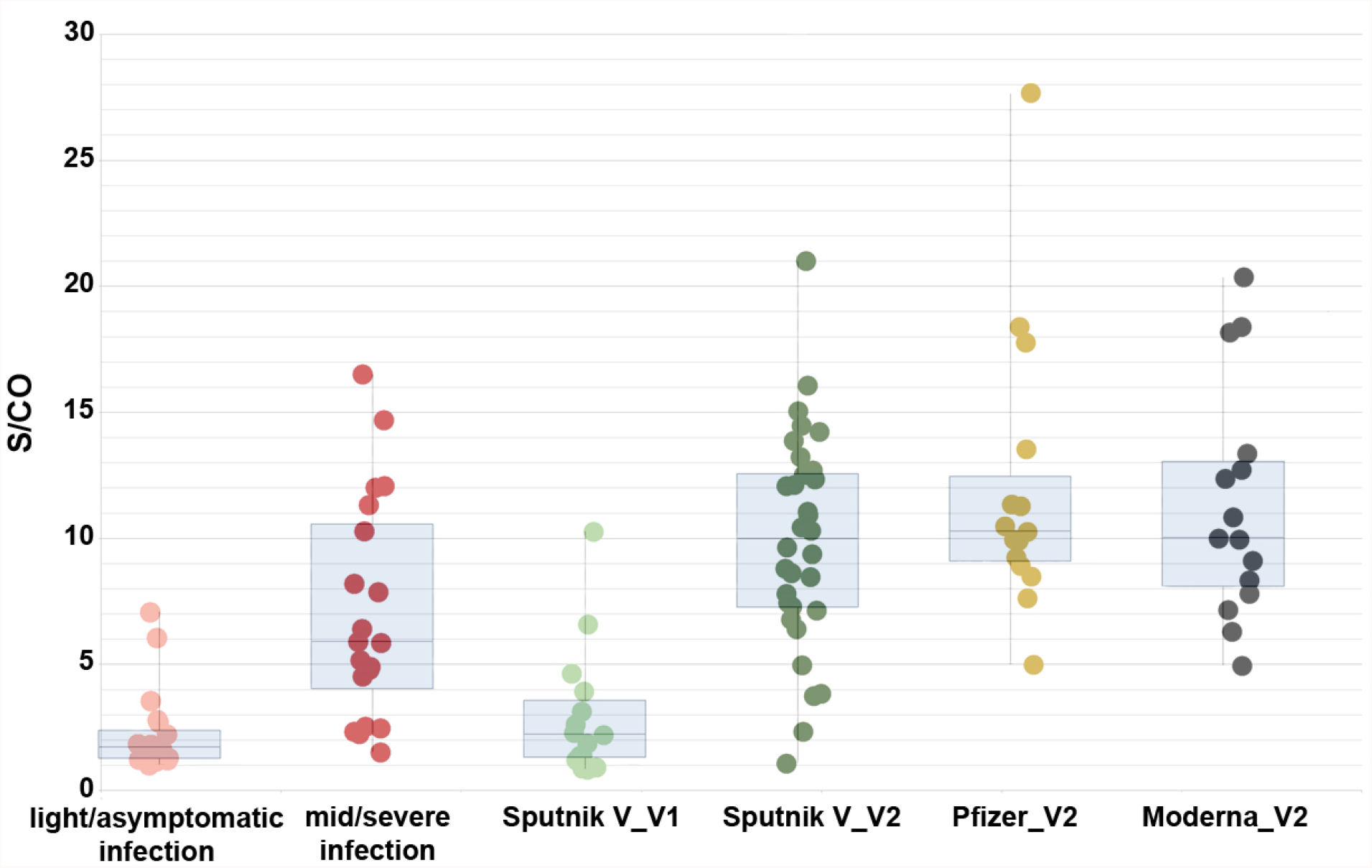
Comparison of S/CO for the anti-RBD IgG antibodies in the individuals after light/asymptomatic course of COVID-19 (pink dots), moderate/severe course (red dots), Sputnik V single dose vaccinated (light green dots), Sputnik V both doses vaccinated (dark green dots), Pfizer vaccinated (yellow dots), and Moderna vaccinated individuals (black dots).

Reasonably, highest levels of antibodies were detected in the individuals with positive S/CO for anti-N antibodies. There were 8 such cases, with respective S/CO for the anti-RBD antibodies ranging from 10.51 to 20.52, with an average of 15.1 and a median of 13.7. We excluded these cases from further analysis here. The 32 remaining Sputnik V two-dose vaccinated samples demonstrated results with an average of 9.53 and a median of 9.51. The record case was a S/CO of 21.0.

For the Pfizer-BioNTech vaccinated individuals, 18 out of 18 cases showed positive anti-RBD antibody results (Fig. 2). 3 cases demonstrated positive results for the anti-N antibodies, and were further excluded (the respective S/CO for the anti-RBD antibodies in these cases were high, 15.73, 12.92, and 13.06). The resulting range was between 4.98 and 27.63, with the average of 11.99 and median 10.27. The record case was a S/CO of 27.63, the highest result obtained in the whole study.

All 16 Moderna-vaccinated individuals were anti-RBD antibody positive, with one further excluded positive anti-N case with the S/CO for anti-RBD antibodies of 23.72. The range was between 4.95 and 20.37, with an average of 11.32 and a median of 10.01 (Fig. 2). The record case was a S/CO of 20.37.

All 40 studied cases of previously infected individuals (who have not been vaccinated) also demonstrated above-threshold results for the RBD antibodies (Fig. 2). The average S/CO was 4.69, ranging from 1.01 to 17.03, with a median of 3.10. The patients in our study consisted of two groups, 20 individuals each: a group with moderate or severe symptoms, and a group with light symptoms or a completely asymptomatic course of the disease. The first group had an average S/CO of 2.22 (ranging from 1.01 to 7.07, with a median of 1.7), and the second group had an average S/CO of 7.08 (ranging from 1.52 to 17.03, with a median of 5.88). As measured by the Mann-Whitney test, we observed a significant difference between these signal distributions (P-value 1.3 × 10^− 05^), in consistency with our previous studies (21).

### 3.3 Antibody signal for the vaccinated groups of individuals of different sex and age

For the 35 fully vaccinated females (both doses of any of the three vaccine types administered, N-positive samples excluded), the average S/CO was 11.37, median 10.08; for the 27 males the average S/CO was 10.98, median 9.91. Although the IgG antibody levels for females were slightly higher on average, the overall distributions of the respective values did not significantly differ (P-value 0.4) (Supplementary Fig. 1).

For the 39 individuals with age less than 50 years the average S/CO was 9.99, median 9.65; for the 22 individuals 50 or older the average S/CO was 13.26, median 10.31. The two distributions of the respective values did not show a significant difference (P-value 0.1). According to Pearson’s Spearman’s coefficient analysis there was no overall correlation between age and RBD signal levels (P-value 0.73).

Similar analysis for each of the three vaccine type groups individually did not demonstrate any significant difference between male/female or younger/older distributions of the S/CO values.

### 3.4 Antibody signal variation with time after full Sputnik V vaccination

To understand if levels of anti-RBD antibodies tend to drop or raise with time within the scope of our study, we have taken additional samples from individuals after the full Sputnik V vaccination, in 54 to 205 days after vaccination (Fig. 3).

**Fig. 3.**
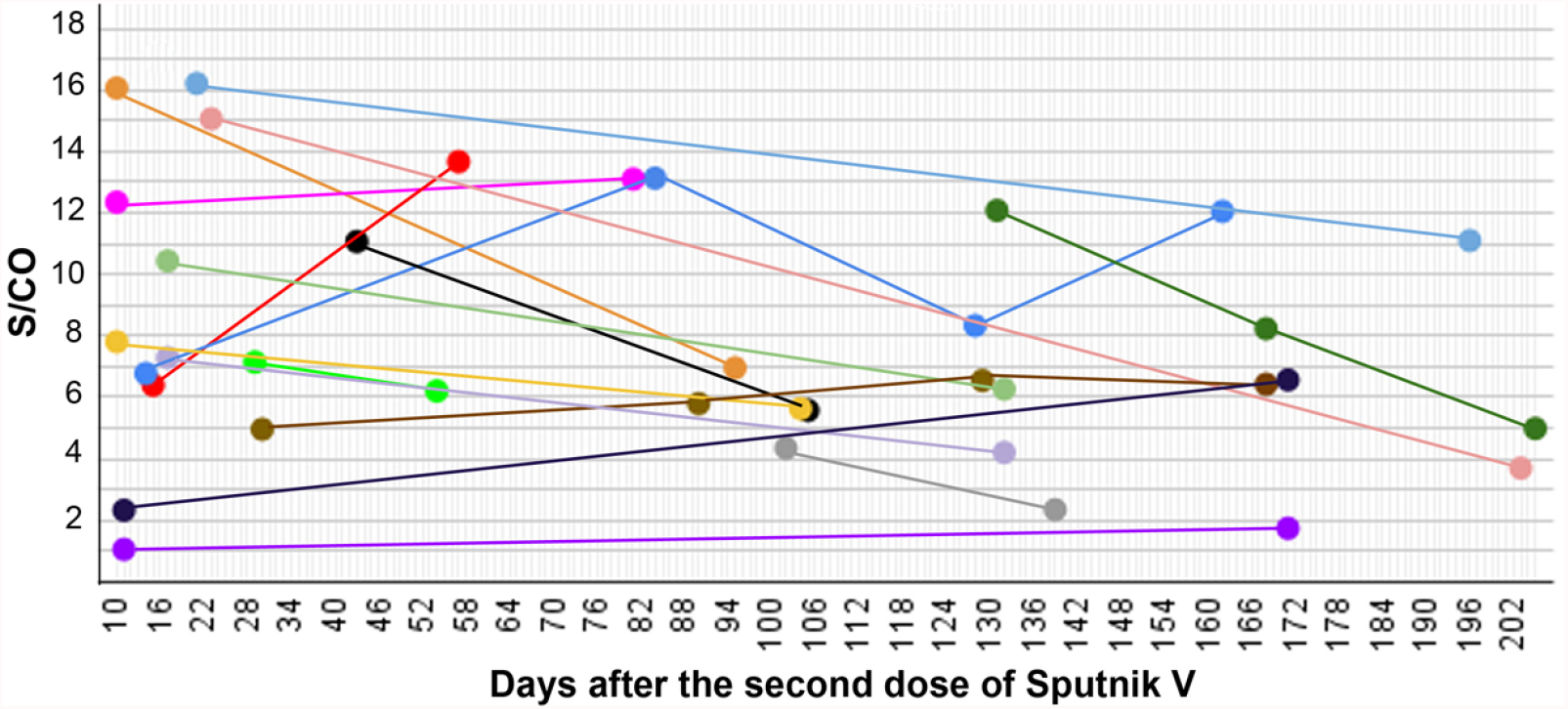
S/CO in 17 individuals measured after full vaccination with Sputnik V at several time points presented as a time course.

Overall, 39 measurements were taken for 17 individuals, at two to four time points for each. In 13 cases the antibody signal dropped over time, and in 9 cases the levels rose. We did not observe a significant correlation between the change in the S/CO and the time between different samplings taken after the vaccination (P-value 0.7).

There was one individual (blue line in Fig. 3) with signal raising by 93% in 70 days, then lowering by 64% in the next 40 days and then, in the next 34 days, raising back by 45%. In another case (red line in Fig. 3), where the anti-RBD antibody level raised by 114%, the anti-N antibody result appeared as positive in the second sampling, most likely indicating an encounter with the SARS-CoV-2 infection in the period of time between the two samplings.

### 3.5. Neutralization effect in samples from the Sputnik V vaccinated and infected individuals

We have assessed the neutralization capabilities of serum after the Sputnik V vaccination using a surrogate virus neutralization system. It allows measuring the effect of antibodies coupling to the viral antigen and preventing it from binding to the human ACE2 receptor. Samples of 30 anti-N-negative individuals who received both doses of Sputnik V were tested. For all vaccinated patients, the neutralization effect was high and varied between 61.59% and 97.74% with an average of 80.69% and median of 80.51% (Fig. 4A).

**Fig. 4.**
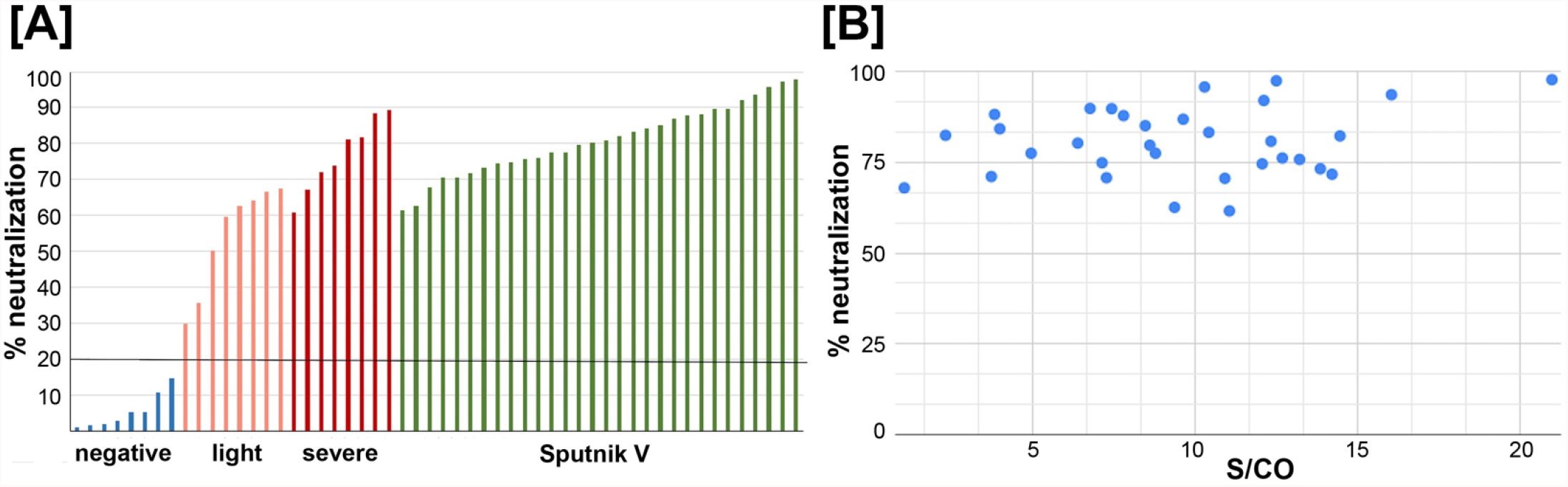
[A] Serum of the Sputnik V vaccinated patients efficiently neutralize interaction of the S-protein RBD domain with ACE2 receptor. Bars for the negative controls representing samples before 2019, are in blue, asymptomatic/light symptom patients are in pink, patients with moderate/severe symptoms are in red, and for the Sputnik V vaccinated individuals are in green. Cut-off level of 20% is indicated by the black horizontal line. N-positive patients were excluded from the analysis. [B] Neutralization effect in samples of both-doses Sputnik V vaccinated individuals. No significant correlation was observed between the effect and the antibody signal.

No significant correlation was observed between the neutralization effect and the antibody signal (P-value 0.2) (Fig. 4B).

In comparison, the neutralization effect measured for patients who had been infected by the SARS-CoV-2 varied greatly, ranging between 39.03% and 89.31%, with the average for the group with no/light symptoms of 58.38% (median 62.02%) and for the group with moderate/severe symptoms of 72.74% (median 73.98) (Fig. 4A), in consistency with our previous studies (21).

### 3.6 Antibody signal and neutralization capabilities for vaccinated individuals previously exposed to COVID-19

As it has been mentioned previously, the observed immune response upon vaccination was higher in patients with positive S/CO for the N antigen. We tested eight such cases, of which four were confirmed with PCR or antigen test during the disease, two claimed to have symptoms of COVID-19 at 94 to 165 days prior to vaccination, and one (PV8) was presumably infected during vaccination (Fig. 5A).

**Fig. 5.**
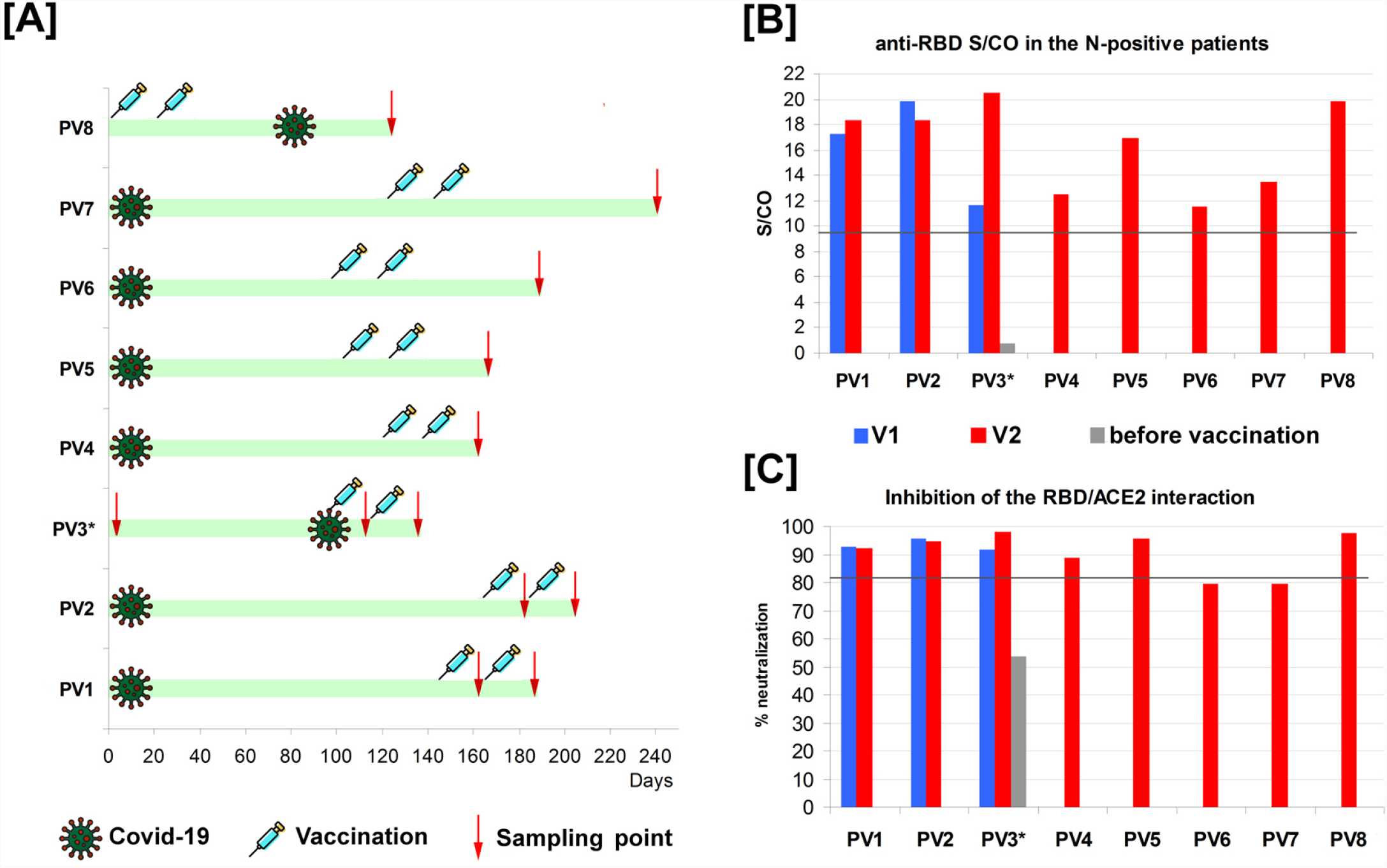
[A] Time points of Covid-19 symptoms, vaccination and sampling. PV3* - no antibodies and no symptoms before vaccination, but severe “side effects” for a week after the first dose [B] Results for S/CO of IgG anti-RBD antibodies in the individuals with positive results for the anti-N antibodies, after first (blue) and second (red) administration of the Sputnik V vaccine. Grey bar is the anti-RBD level before vaccination. Threshold S/CO value is 1. [C] Inhibition of the RBD interaction with ACE2 in the individuals with positive results for the anti-N antibodies, after the first (blue) and the second (red) administration of the Sputnik V vaccine. Grey bar is the neutralization level before vaccination.

In the last case, PV3*, patient was sampled several times, 150, 120, and 95 days before vaccination, with stable negative results for N-antigen in all cases. No symptoms were reported before the vaccination. After vaccination, the patient developed severe side effects, with high fever above 38°C for almost a week, possibly indicating infection with SARS-CoV-2 during vaccination or just before it. In line with this assumption, their respective levels of anti-N antibodies measured after the second dose (5 weeks after possible infection) slightly increased, while the anti-RBD levels increased almost 2-fold (Fig. 5B).

We also measured the antibody levels before and after the second dose for two N-positive patients with confirmed infection. Neither of them demonstrated significant change in the anti-RBD S/CO after the second administration of the vaccine.

Overall, the respective results for N-positive patients were among the highest values obtained for the anti-RBD antibody signals, exceeding the threshold 12-20 times (15.13 times on average, median 13.5) (Fig. 1 and 5B). The difference between the distributions of S/CO of the antibodies to the anti-RBD antigen within N-positive and N-negative samples as measured by Mann-Whitney test was very prominent (P-value 0.0001).

The same tendency was observed for the neutralization capability of respective serum (Fig. 5C); 6 out of 8 samples were above the calculated median for the N-negative vaccinated individuals, and almost no changes were detected after the second vaccine administration.

## Discussion

It has been shown in various studies that Moderna, Pfizer-BioNTech, and Sputnik V vaccines are capable of producing a prominent immune system response, yielding vast development of the anti-spike protein antibodies (11,22,23). However, it was of interest to analyze the respective signals using a unified testing system which would allow us to compare them directly. In this study, we have used the VirIntel dual-antigen assay, an ELISA-based system, testing serum simultaneously for the IgG antibodies to the nucleocapsid and the receptor-binding domain of the spike protein (20). This, in particular, allowed us to separate cases of individuals possibly infected by SARS-CoV-2 before the vaccination, who had antibodies to the N-antigen. Our first goal was to compare the anti-RBD IgG antibody levels in samples obtained from the three vaccinated groups. Sex and age of individuals were then assessed as possible factors affecting the antibody levels. Next, we compared vaccinated individuals with two groups of patients previously infected by SARS-CoV-2, with light/asymptomatic or moderate/severe course of the disease. We also assessed the neutralization capabilities of anti-RBD antibodies in Sputnik V vaccinated individuals and whether their antibody levels tend to drop or rise within several weeks after vaccination. Finally, the antibody levels and neutralization capabilities in several individuals who have been infected and vaccinated were studied.

### 4.1 Anti-RBD antibodies signal levels are similar in Moderna, Pfizer-BioNTech, and Sputnik V vaccinated individuals

All three studied vaccines demonstrated a prominent immune system response, yielding the production of IgG antibodies to the viral RBD antigen. All of them require two-dose administration. We first compared the levels of anti-RBD IgG antibodies in the individuals three weeks after the first dose and two or three weeks after the second dose of Sputnik V. After a single dose of Sputnik V only 11 out of 14 individuals demonstrated positive results. After the second dose, all the individuals demonstrated positive results. This highlights the importance of the two-stage vaccination of naive patients.

The signal levels after the second dose of Sputnik V were significantly higher, on average 5.4 fold. At the same time, we did not observe significant correlation between the signals for first and second sampling within individuals, for some of them the signal raised only slightly, for others more than 7-10 fold. This might reflect the preexisting immunity of respective individuals to either Ad26 or Ad5 (two strains of adenoviruses used for the first and second dose of Sputnik V, respectively)

For individuals vaccinated by both doses of Sputnik V, Moderna or Pfizer-BioNTech a wide range of signals was observed in each group, going up to values exceeding threshold over 20 times, with the Pfizer-BioNTech vaccine showing highest values. These ranges most likely reflect specific features of the immune system of different individuals.

At the same time, the average and median S/CO values of all three groups were similar (Table 1). Moreover, distributions of these values did not demonstrate significant difference (P-value above 0.05 for each possible pair of groups). This finding indicates that all three vaccines not only induce a strong immune system response, but are also comparable with each other. They yield similar levels of anti-RBD IgG antibody production, despite the difference in respective vaccine structures, Moderna and Pfizer-BioNTech being RNA-based vaccines, and Sputnik V an adenovirus vector.

**Table 1.** Signal/Cut-off ranges, average and median values of the analyzed sets of samples from vaccinated and previously infected individuals.

| Vaccine/Course of disease | Range of S/CO | Average S/CO | Median S/CO |
| --- | --- | --- | --- |
| Sputnik V | 1.06 - 21.0 | 9.53 | 9.51 |
| Pfizer-BioNTech | 0.83 - 27.63 | 11.99 | 10.27 |
| Moderna | 1.07 - 17.78 | 11.32 | 10.01 |
| Light/asymptomatic disease | 1.01 - 7.07 | 2.22 | 1.7 |
| Moderate/severe disease | 1.52 - 17.03 | 7.08 | 5.88 |

### 4.2 Sex and age do not seem to affect anti-RBD antibody signals in vaccinated individuals

Reports indicate that the number of COVID-19 cases between men and women is similar, but men experience more severe outcomes, including hospitalization, admission to the intensive care unit, and death, the difference in risks being sometimes estimated as two-fold (24). On the other hand, females have been reported to have higher risks of acquiring side effects after vaccination against COVID-19 (25).

It was thus of interest to compare the levels of anti-RBD IgG antibody development in males and females after vaccination. This type of studies have been done for other vaccines; in most cases females have been shown to have greater immune system response (26,27), in particular, for the influenza vaccine (28), some studies even suggest half-dose vaccination for females to be more adequate (29). The vaccines typically administered to children, such as combination measles, mumps, and rubella (MMR) vaccine also elicited higher antibody development levels in girls (30).

In our study, for the 35 studied fully vaccinated females, the antibody level was on average only 3.5% higher than for the 27 studied males. The distributions of the respective values did not demonstrate a statistically significant difference, thus we did not observe a correlation between sex and the levels of the anti-RBD IgG antibodies of vaccinated individuals.

Aging can also be a significant factor in the immune system response after different types of vaccinations, in particular, reducing the antibody development levels (28,31–33). SARS-CoV-2 anti-spike IgG antibody levels have been recently shown to be lower for people over the age of 80 vaccinated by the Pfizer-BioNTech vaccine than for people under the age of 60 (33). However, the subject of vaccine-induced immunity correlation with age still remains unclear; other studies have reported that the vaccine efficacy is similar for younger and older cohorts of individuals. In particular, during the clinical trials of the Pfizer-BioNTech vaccine, breaking down the participants by age revealed that of 3,848 vaccine recipients older than 65 years of age only one became infected whereas 19 of 3,880 placebo recipients of that age tested positive. This translated to an estimated efficacy of 94.7%, highly similar with the 95.6% in those aged 16–55 and the 93.7% in those aged 55–65 (34).

In our study, for the 23 fully vaccinated individuals with the age of over 50 the average signal was 14% higher than in the older group of 39, but the two distributions of values did not demonstrate a significant difference (P-value 0.1). Overall correlation between the signal and age values was also not observed (P-value 0.9). In particular, several samples taken from people over the age of 70 yielded results 8-16 times exceeding the threshold value, on a par with people of around 50 or 35 years old. Only one vaccinated individual of over 80 was present in our dataset (vaccinated by Moderna), and their respective anti-RBD S/CO was 6.3, which was, in concordance with the aforementioned study, on the lower scale of the dataset values. At the same time, an over 70 years old patient vaccinated by Sputnik V demonstrated anti-RBD S/CO of 16.06 which is in the upper scale of all results. For future studies, more samples of individuals within the older age group would be required.

### 4.3 Anti-RBD antibody development are similar in vaccinated individuals and previously infected patients with moderate or severe course of the disease

It has been shown in previous studies that patients with severe COVID-19 symptoms generally develop more antibodies to the virus (35,36). In accordance with this, we have shown significant difference between antibody signal distributions for the two studied groups: with asymptomatic/light course of the SARS-CoV-2 infection and with moderate/severe symptoms, both in this and our previous studies (21).

Further analyzing the IgG anti-RBD results for the group of individuals with moderate/severe symptoms, we have shown that the respective distribution of obtained values also did not significantly differ from any of the studied vaccinated groups. On the other hand, each of the three vaccinated groups demonstrated significant difference with the distribution of values from the group with an asymptomatic/light course of the disease (P-value 3.4e-08, 7.3e-05 and 0.001 for Sputnik V, Moderna and Pfizer-BioNTech vaccinated groups, respectively). These results indicate that the general trend of the development of the IgG antibodies is similar for vaccinated individuals and for previously infected patients, but only with prominent symptoms of COVID-19.

### 4.4 Antibody level do not necessarily lower within many weeks after full Sputnik V vaccination

Studies about the longevity of the antibody levels to SARS-CoV-2 are not in full agreement, some reporting rapid waning of virus-specific IgG antibodies by several months after the infection (37,38), and others observing stable titers detected over the same periods of time (39–41). In a study on COVID-19 patients with different courses of the disease including death cases, it was suggested that quality rather than quantity of antibodies may predict the outcome of the infection (42). On the other hand, low or undetectable titers of antibodies are currently a reason for recommending vaccination by doctors in many countries.

It was recently shown that nonhuman primates infected with SARS-CoV-2 after vaccination with spike-based DNA vaccines yielded strong immune responses, including neutralizing antibodies, which possibly indicates that antibody development may in some cases be more effective in preventing than resolving the disease (43). Studies of the mRNA vaccines in humans have demonstrated that antibody levels after the administration seem to be detectable and persistent within several month periods (44,45), although an overall decline in respective levels has been reported. On the other hand, stable titers were reported for the adenovirus-based ChAdOx1 vaccine Oxford–AstraZeneca (46). It was thus of interest to assess the changes in antibody levels in the adenovirus-based Sputnik V vaccinated individuals.

Analysis of the serum of 17 Sputnik V fully vaccinated individuals in 7-30 weeks after the initial sampling demonstrated that anti-RBD levels do not have an obvious tendency towards lowering with time. In 13 cases the signal dropped, and in 9 cases it rose, and there was no significant correlation between time and signal change (P-value 0.7). In the cases where more than two samplings were taken, all possible scenarios were played out, with signal raising, dropping or first dropping and then raising again and vice versa.

Such variations might reflect individual aspects of one’s immune system or point to an encounter with the SARS-CoV-2 at uncertain time points between samplings. In the latter case, the rise of the anti-RBD antibody signal may indicate the respective immune response upon interacting with the virus and quickly conquering it. In one studied case the anti-nucleocapsid antibody result was positive along with the significant rise of the anti-RBD antibody level (by 113.5%). This most likely indicates that the respective individual unknowingly encountered SARS-CoV-2 infection between the two samplings and here, unlike the other cases, this interaction was long enough for their immune system to produce other kinds of antibodies as well.

Further analysis should include more samples and time points, but these results indicate that the stability of the anti-RBD IgG antibody levels over time after the Sputnik V vaccination is not easily predictable, and most likely depends on the individual characteristics of the immune system of a person. At the same time, the S/CO stayed positive for all studied individuals, for the vast majority of samples exceeding the threshold level over four times even after more than a half year since the vaccination. This points to prominent persistence characteristics of the adenovirus-based vector vaccine Sputnik V in the studied period of time.

### 4.5 Sputnik V vaccine effect on previously infected individuals

It has been recently shown that the IgG antibody titers of vaccinated individuals with preexisting immunity can be 10 to 45 times as high as those without it (47), even after administration of a single dose of the mRNA vaccines (45). It was also shown that the second dose does not significantly affect the level of the antibodies within the same individual (47). Moreover, a single immunization boosted neutralizing titers of respective antibodies by up to 1000-fold (48).

In this study we demonstrate that the adenovirus-based Sputnik V yielded similar results. We analyzed 11 samples of 8 vaccinated individuals, which showed positive results for the anti-nucleocapsid antibodies (3 samples were taken after the first dose of the vaccine, 8 after the second). The signal to cut-off ratio values for the N-positive samples taken after the first dose were all quite high (average 15.58), while the signals for the N-negative individuals after the first dose were on average 4.5 times lower. The S/CO values for the N-positive individuals after the second dose of Sputnik V were on average 15.14, thus not showing significant difference compared to the first dose group (the average for the respective N-negative samples was 1.6 times lower).

Overall, we show that individuals who had been previously exposed to COVID-19 demonstrate very high levels of antibodies after the vaccination with Sputnik V, in concordance with the results obtained in other studies for Moderna and Pfizer-BioNTech vaccines. This can be explained as a strong response of the immune system, which most likely treats the vaccine at least partially as a SARS-CoV-2 reinfection and yields vast antibody production. Our results indicate that a single administration of the Sputnik V (Sputnik Light) could be a sufficient boost to the immune system for those who have had the SARS-CoV-2 infection, that is in line with other studies on Sputnik Light (47,49–51).

### 4.6 Neutralization effects are high for Sputnik V vaccinated and are comparable with previously infected individuals with moderate and severe course of COVID-19

SARS-CoV-2 integrates in the human cells, at least partially, via the interaction of the viral RBD S-domain with the ACE2 receptors on the cell surface. Respective neutralizing effects, in turn, are shown to be highly predictive in terms of protection against symptomatic course of the COVID-19 (52). Thus, one of the expected outcomes of vaccination should be the ability of serum to efficiently prevent this interaction. Such neutralization effects were demonstrated, in particular, for individuals vaccinated by mRNA-based vaccines (53,54). In the Sputnik V clinical trials, the seroconversion level in the microneutralization assays with tissue cultures was reported to be 95.38% (55). In (56), eleven candidate vaccines were compared from the viewpoint of peak neutralizing antibody response, and according to the data available at that moment, the highest production of neutralizing antibodies were induced by BBIBP-CorV, AZD1222 and BNT162b2, followed by New Crown COVID-19 and Sputnik V. In line with this, we observed a high capability of serum from the Sputnik V vaccinated individuals to inhibit the interaction of RBD with ACE2. We have found that the average rate of the inhibition was 81%, which was significantly higher than in the patients with light symptoms (58%), and closer to that in patients with a moderate/severe course of COVID-19 (72%). This is in concordance to previous observations for mRNA vaccines (56–58).

Similar to the anti-RBD IgG levels, neutralization effects were very prominent in the vaccinated individuals with previous SARS-CoV-2 infection, with a record of 98.02%. In those cases, high neutralization rates were observed after the first dose of Sputnik V and did not significantly change after the second. This supports recent proposals for application of a single dose vaccine boost for previously infected patients (47,59).

## Supporting information

Supplemental Fifure 1

## Data Availability

All data produced in the present work are contained in the manuscript

## Acknowledgements

We thank the Laboratory of cell cultures and cell engineering of the Institute of Cell Biophysics RAS for providing pre-pandemic samples, and Dr. Oleg S. Morenkov for useful suggestions throughout this project. We also thank Denis Kaznadzey for help with experiment logistics and for fruitful discussions.

## Conflicts of interest

Authors (I.M., A.K., M.K.) are employees and consultants of VirIntel, LLC, the company which has developed the testing system used in the article. Authors are inventors of a pending patent on this testing system. Author IM is also employed by Argentys Informatics, LLC. The remaining authors declare that the research was conducted in the absence of any commercial or financial relationships that could be construed as a potential conflict of interest.

## Author contributions

AK and MT have contributed equally to this work and share first authorship. MK, MT and TB performed experiments. AK performed statistical analysis. AK and MT wrote the article. AK, MT, MK, and IM reviewed the article. IM supervised the study and obtained funding. All authors approved the submitted version.

## Funding

This research received no external funding, it was funded by internal company funds.

## Notes

### Author Declarations

All procedures were approved by the Commission on Biosafety and Bioethics of the Institute of Cell Biophysics, Pushchino Scientific Center for Biological Research of the Russian Academy of Sciences. All participants provided their written informed consent to participate in this study.

