## Supplementary figures and images for "BNT162b2, mRNA-1273, and Sputnik V vaccines induce comparable immune responses on a par with severe course of COVID-19"

### Supplemental Fifure 1

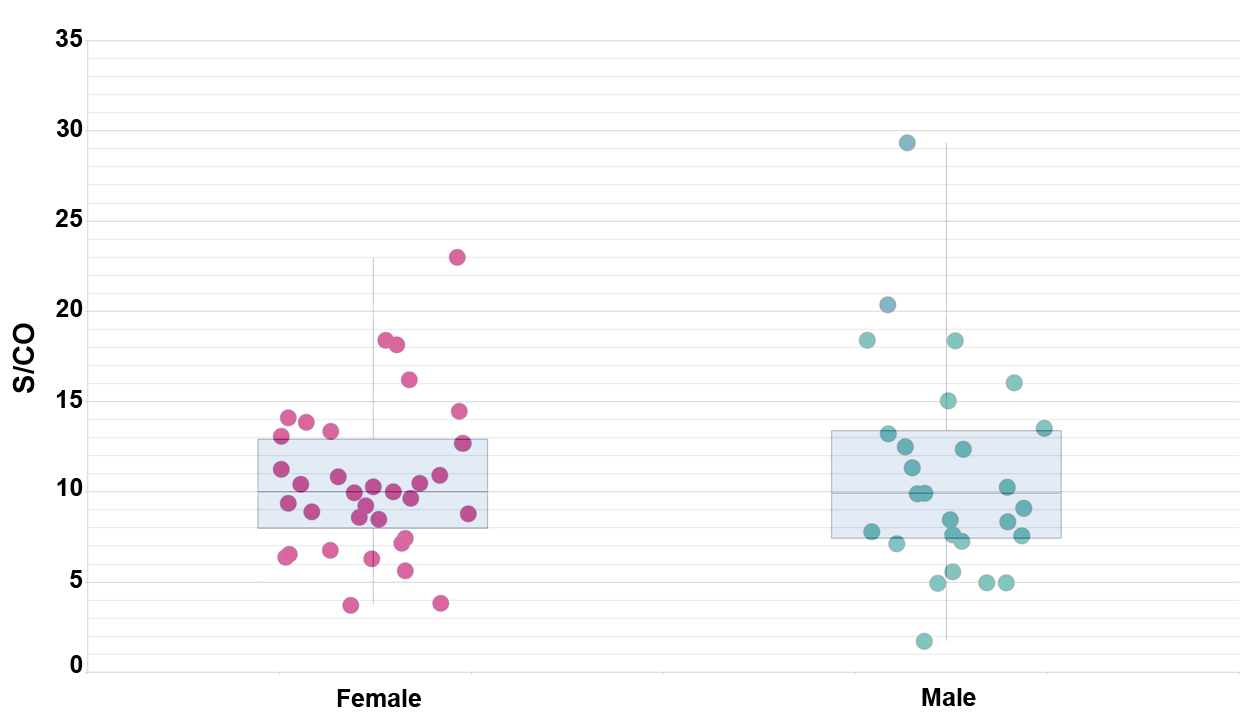
